# Does chronotype influence the odds of diabetes comorbidity in bipolar disorder? A cross-sectional analysis of the Our Future Health cohort

**DOI:** 10.64898/2026.09.29.26364301

**Authors:** Duncan Swiffen, Arish Mudra Rakshasa-Loots, Christina Steyn, Haya Deeb, Kelly Fleetwood, Cathy Wyse, Daniel J. Smith

## Abstract

**Background:** Diabetes is common in people with bipolar disorder, particularly men. Both type 2 diabetes and bipolar disorder are associated with evening chronotype, and men of working-age have later chronotypes than women. However, it is not known whether elevated rates of diabetes in bipolar are explained by evening chronotype, nor whether sex differences in chronotype contribute to higher rates of diabetes observed in men with bipolar disorder.

**Aim:** To determine whether evening chronotype increases the likelihood of comorbid type 2 diabetes in bipolar disorder, relative to those with no mental health problems. Additionally, to determine whether any effect differs between men and women.

**Methods:** Using data from the Our Future Health cohort, we estimated odds ratios for type 2 diabetes by bipolar disorder diagnosis, chronotype, and their interaction, adjusting for relevant covariates.

**Results:** A total of 1,136,694 participants (N = 6,846 with bipolar disorder) were included in the adjusted models. Type 2 diabetes was independently associated with bipolar disorder and evening chronotype. There were no significant differences in the interaction effects of chronotype on the odds of type 2 diabetes in bipolar across the whole sample or when stratified by sex.

**Conclusions:** The high prevalence of type 2 diabetes in people (and in men, specifically) with bipolar disorder may be independent of evening chronotype.

## Introduction

Bipolar disorder is strongly associated with circadian dysregulation (Zimerman et al., 2025). Sleep disruption, for example, is both a symptom of and a trigger for mood episodes in bipolar, and can act as an early warning sign for an emerging mood episode (Bipolar UK, 2025).

Chronotype – a preference for activity at a particular time of day – is also associated with bipolar disorder, with evening chronotype being more common than in the general population (Romo-Nava et al., 2020). This may have implications for the health of people with bipolar more generally, as evening chronotype is associated with higher rates of cardiometabolic disorders, including type 2 diabetes (Heikkinen et al., 2025). Previous studies have identified that men between the ages of 16 and 50 tend to have later chronotypes than women (Walton et al., 2022), potentially further increasing their risk of cardiometabolic disorders.

Using data from the world’s largest consented research cohort (Our Future Health, 2025), we previously found that people with bipolar disorder have higher likelihood of cardiometabolic comorbidity compared to those with no mental health problems (Swiffen et al., preprint). We also observed a higher prevalence of cardiometabolic comorbidity in males than in females with bipolar disorder. The aim of this study is to investigate the relationship between chronotype, sex, bipolar disorder and type 2 diabetes. Specifically, we assess whether chronotype modifies the association between bipolar disorder and type 2 diabetes and, if so, whether this differs between males and females.

## Materials and Methods

### Our Future Health

The Our Future Health (OFH) cohort comprises adults aged 18 years or older living in the UK. Recruitment to OFH is described elsewhere (Swiffen et al., preprint). All participants provided electronic informed consent at baseline. Ethical approval for OFH was acquired from the Cambridge East NHS Research Ethics Committee (ref: 21/EE/0016).

Our analysis was conducted using Data Release 14 (v14.0+daf23ff) as part of approved study ID OFHS240114. Data from OFH is accessible through a Trusted Research Environment supplied by DNAnexus, which can be accessed by authorised researchers named on specific projects approved by the OFH Access Board.

Responsibility for interpretation of the information supplied by OFH is the authors’ alone.

### Study design

The current cross-sectional analysis used data from the OFH baseline questionnaire (version 2.2, November 2022), data collected at in-person clinic appointments, and geographic data in the form of Lower layer Super Output Areas (LSOA) (Ministry of Housing Communities & Local Government, 2019).

We used questionnaire data to categorise participants into two mental health groups – i) Bipolar disorder, and ii) No mental health problems – and four chronotype levels – i) ‘Definite Morning’, ii) ‘More Morning than Evening’, iii) ‘More Evening than Morning’ and iv) ‘Definite Evening’. We excluded participants with invalid answers to the relevant questions, and participants who only self-reported a mental health problem other than bipolar disorder.

We removed participants who completed version 1 of the questionnaire due to non-comparable wording of relevant questions. We also excluded participants reporting night shift work, given its disruptive effect on circadian rhythm, and participants who reported an absence of mental health problems despite reporting taking medications for their mental health (other than sleeping medications).

Our outcome measure, a diagnosis of type 2 diabetes, was self-reported as part of the baseline questionnaire.

Participants with data missing for any covariate were removed from analysis. LSOA data in OFH is currently available for England-based participants only, so our sample does not include participants living elsewhere in the United Kingdom.

Detailed description of variables used in our analysis is available in Supplementary Materials.

### Statistical analysis

We used logistic regression to estimate odds ratios and 95% confidence intervals for the association of bipolar disorder and chronotype with type 2 diabetes. We fitted two models, the first included bipolar disorder (versus no mental health problem) and chronotype (versus a ‘definite morning’ chronotype), and the second model additionally included a term for their interaction. We adjusted both models for age, sex, ethnicity, smoking status, current alcohol use, educational attainment, Index of Multiple Deprivation (IMD), physical activity levels, body mass index (BMI), use of sleeping medications, and multimorbidity score. Age and BMI were modelled continuously using both linear and quadratic terms; all other covariates were categorical.

The same models were then repeated on the sample stratified by sex (sex as a covariate was removed in these models).

To determine whether the association between bipolar disorder and type 2 diabetes differed by chronotype, we conducted likelihood ratio tests for the whole sample and in sex-stratified analyses to compare nested regression models with and without the interaction term. P-values were adjusted for multiple comparisons using the false discovery rate (FDR) correction.

The data was analysed using R v4.4.0. Analysis code is available through the following link:

<https://github.com/DSwiffen/OFH-ChronotypeDiabetesBipolar-August2026.git>

## Results

### Sample

Of the 2,024,465 participants who completed baseline questionnaires, we included 1,280,610 participants after exclusions (Supplementary Figure 1). The most common exclusion was an inability to categorise into either mental health group, followed by an invalid or missing chronotype response. Missingness in at least one covariate further reduced the sample to 1,136,694 participants for fully adjusted logistic regression models. Complete and incomplete cases are compared in Supplementary Table 1.

### Baseline characteristics

Compared to those without mental health problems, participants with bipolar disorder were younger and more likely to be female, have an evening chronotype, a history of regular smoking, and a higher BMI with lower physical activity, but had lower current alcohol consumption. They were less likely to have a college or university degree and had greater socioeconomic deprivation and more physical comorbidities (Table 1).

Of those with bipolar disorder, males were older, and more likely to have a history of regular smoking and frequent alcohol use, but were more physically active, with fewer self-reported physical comorbidities and less use of sleeping medications compared to females.

### Type 2 diabetes in bipolar disorder and across chronotypes

Type 2 diabetes was most prevalent among males with bipolar disorder (7.9%) and least prevalent among females with no mental health problems (1.6%) (Table 1).

In the model excluding the interaction term, participants with bipolar disorder had higher odds of type 2 diabetes than those with no mental health problems, independent of chronotype (adjusted odds ratio, OR [95% confidence interval; p-value]: 2.21 [1.98, 2.47; <0.001]) (Fig. 1a). Compared with ‘Definite Morning’ chronotype, evening chronotypes had higher odds of type 2 diabetes, independent of mental health status (‘Definite Evening’ OR 1.44 [1.37, 1.50]; <0.001; ‘More Evening than Morning’ OR 1.19 [1.15, 1.23]; <0.001) (Fig. 1b).

**Table 1.**
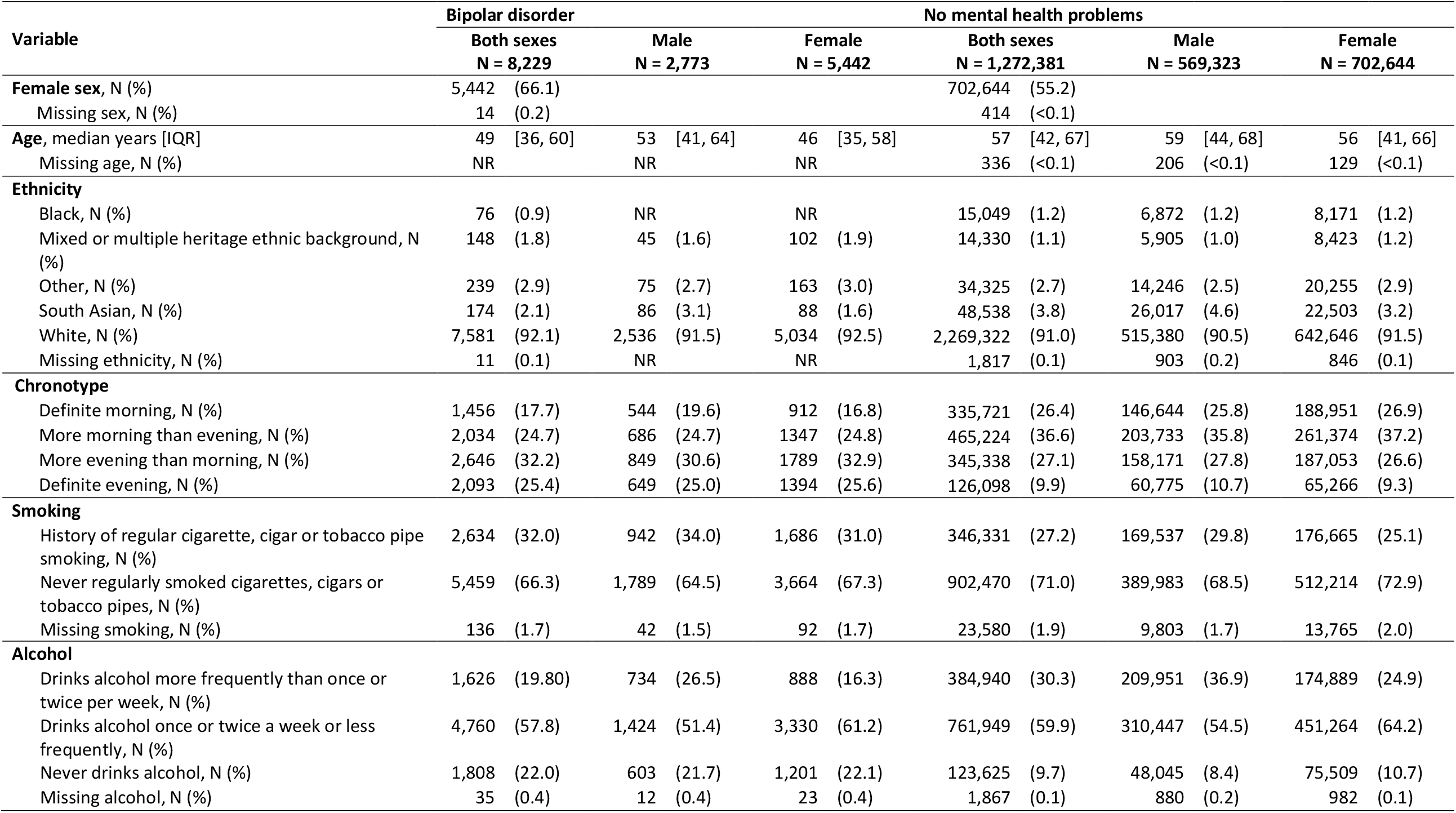
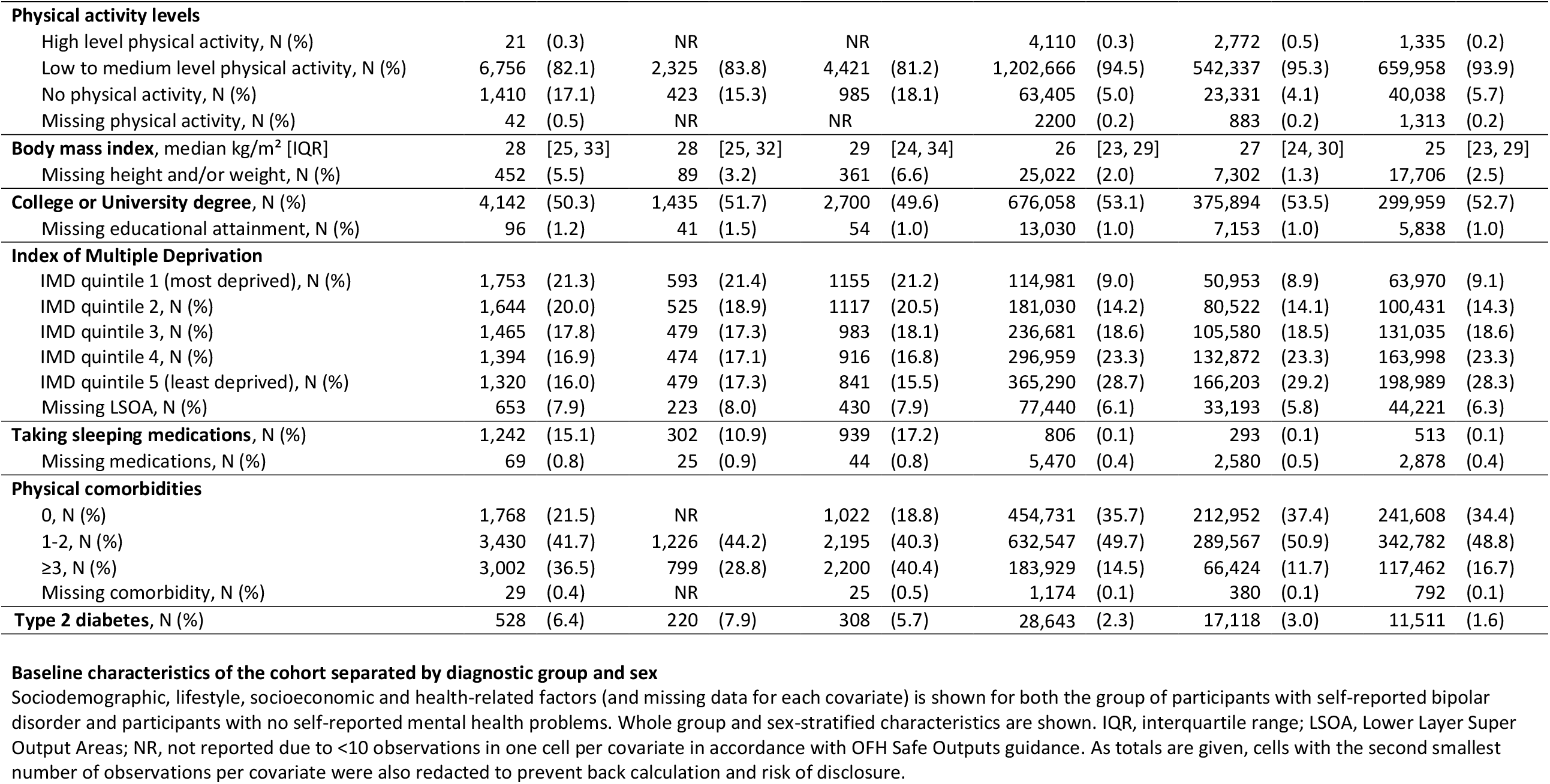

**Figure 1:**
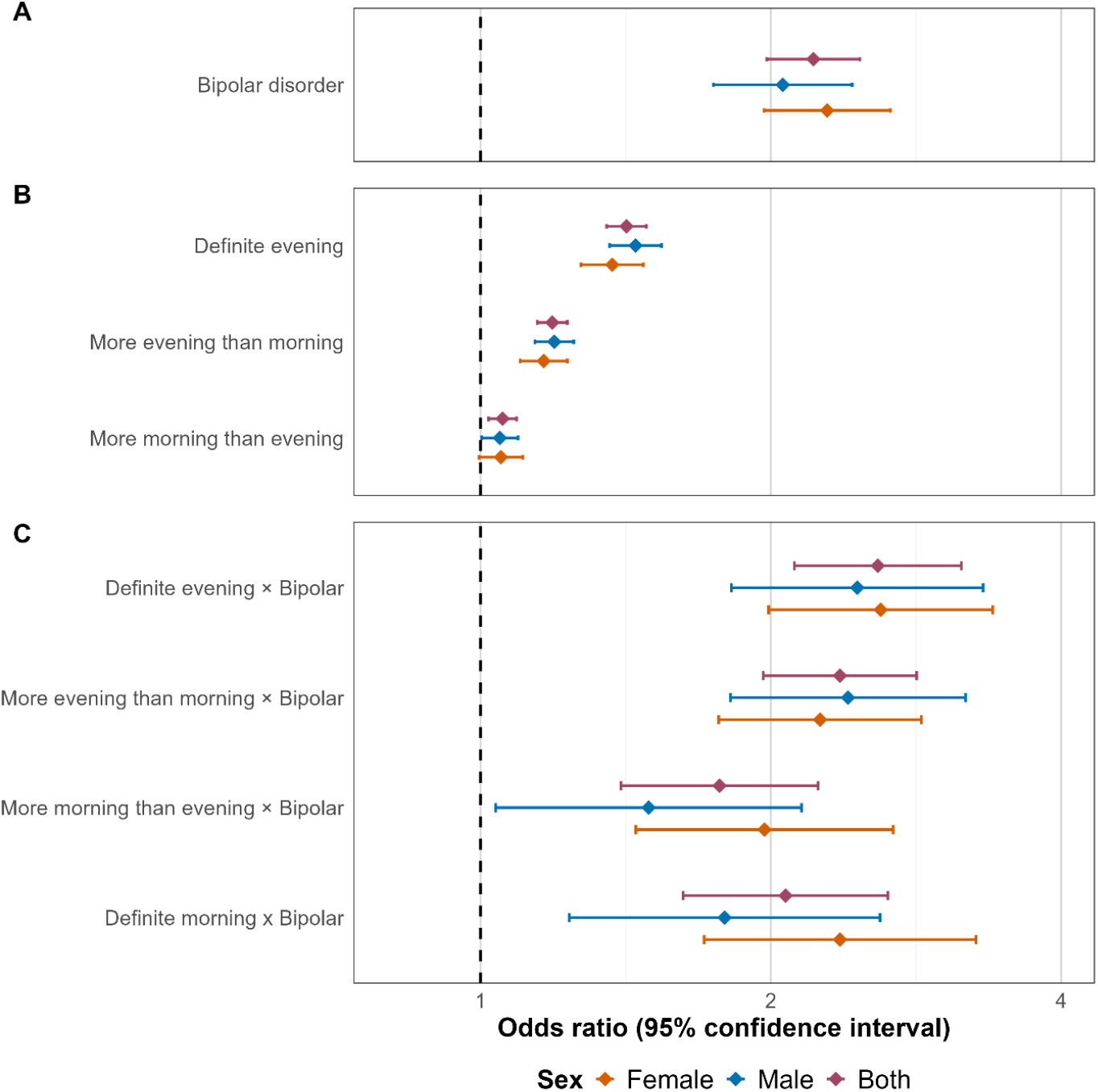
Whole-sample and sex-stratified adjusted odds ratios of type 2 diabetes in those with bipolar disorder versus those with no mental health problems independent of chronotype (**A**), in those with different self-reported chronotype versus a ‘Definite Morning’ chronotype independent of mental health status (**B**), and when estimating the association between bipolar disorder and type 2 diabetes separately within each chronotype (**C**). Odds ratios are adjusted for age, sex, ethnicity, smoking status, current alcohol use, educational attainment, index of multiple deprivation (IMD), physical activity levels, body mass index (BMI), use of sleeping medications, and physical multimorbidity score.

### Interaction between chronotype and bipolar disorder

When including the interaction term, there was no evidence that chronotype modified the association between bipolar disorder and type 2 diabetes (likelihood ratio test, FDR-corrected p = 0.18). Compared to participants with no mental health problems, those with bipolar disorder had approximately twofold higher odds of type 2 diabetes across all chronotypes: ‘Definite Morning’ OR 2.09 [1.64, 2.67], ‘More Morning than Evening’ OR 1.78 [1.41, 2.25], ‘More Evening than Morning’ OR 2.36 [1.96, 2.83], and ‘Definite Evening’ OR 2.53 [2.08, 3.09] (Fig. 1c). Although odds ratios were numerically higher in evening chronotypes, the difference was not statistically significant.

### Sex-stratified interaction analysis

Inclusion of the interaction between bipolar and chronotype did not improve model fit in females (FDR-corrected p = 0.63) or males (FDR-corrected p = 0.18), indicating that the association between bipolar disorder and type 2 diabetes did not differ by chronotype in either sex.

## Discussion

Using data from the world’s largest consented cohort, we found no evidence that the association between bipolar disorder and type 2 diabetes differed by chronotype.

Our finding that type 2 diabetes odds were over twofold higher in bipolar disorder is consistent with previous studies (Breznoscakova & Pallayova, 2022) and reinforces the importance of routine metabolic monitoring in this patient group. Evening chronotype was also associated with type 2 diabetes independent of mental health status, consistent with previous findings (Lotti et al., 2022), suggesting chronotype may be a marker of metabolic risk beyond psychiatric populations. This highlights circadian health as a potential target for optimising metabolic health, although prospective interventional studies are needed to determine whether modifying circadian behaviours reduces diabetes risk.

As both bipolar disorder and evening chronotype are associated with metabolic dysregulation (Breznoscakova & Pallayova, 2022; Heikkinen et al., 2025), we anticipated that that their co-occurrence would confer a compounded risk of type 2 diabetes. There are several plausible reasons why we did not see such an effect. Firstly, there may be a genuine absence of interaction and bipolar disorder and evening chronotype may have additive rather synergistic effects on the odds of type 2 diabetes. It is also possible that bipolar disorder and evening chronotype influence odds of type 2 diabetes through shared biological or behavioural pathways, limiting any additional modifying effect of chronotype. For example, chronic sleep disturbance and inflammation, which are features of both bipolar disorder and evening chronotypes, are known to impair insulin sensitivity and glucose homeostasis (Breznoscakova & Pallayova, 2022; Marchetti et al., 2025; Merikanto et al., 2012; Spiegel et al., 2005) and may represent overlapping mechanisms through which both exposures increase diabetes risk. Alternatively, other factors that we could not account for in our analysis, such as psychotropic medications, may outweigh the comparatively modest contribution of chronotype to diabetes risk, limiting our ability to detect effect modification.

We additionally found no between-sex differences in the odds of type 2 diabetes across chronotypes in bipolar disorder. However, this may have been missed due to the older age of the OFH cohort (Our Future Health, 2025). Men generally exhibit later chronotypes than women, although this sex difference tends to diminish after age 50 years (Walton et al., 2022). Repeat analysis in a younger population would ensure that these findings are robust across all ages.

Our findings emphasise the need to investigate markers of circadian disruption beyond chronotype alone. For example, social jetlag, arising from misalignment between an individual’s circadian and social timing (Taillard et al., 2021), may be particularly relevant to metabolic health, as meal timing plays an important role in body weight regulation and glucose metabolism (Reytor-González et al., 2025), and restricting food intake to specific times of day may confer metabolic benefits (Chen et al., 2026). Given their higher prevalence of evening chronotype, people with bipolar disorder may be especially susceptible to social jetlag, disrupted meal timing, and the resulting metabolic consequences. Ongoing studies evaluating time-restricted eating in bipolar disorder (Johnson et al., 2024) may therefore provide valuable insights into whether targeting circadian alignment can improve metabolic health in this population.

A key strength of this study is the scale of the Our Future Health cohort, which includes the largest bipolar disorder subsample of any UK-based consented cohort (Smith et al., 2013), providing sufficient statistical power to adjust for a broad range of covariates. Nevertheless, several limitations should be considered. The cross-sectional design precludes causal inference, and all measures, including chronotype and diagnoses of bipolar disorder and type 2 diabetes were self-reported, potentially introducing misclassification. Chronotype was assessed using a single-item measure, consistent with previous studies (Romo-Nava et al., 2020), although the absence of an intermediate response category may have contributed to the high level of missingness (Montaruli et al., 2021). In addition, potentially important confounders such as meal timing and dietary composition were unavailable. Future studies should incorporate objective measures of circadian function, such those derived from actigraphy (Geoffroy et al., 2015), alongside longitudinal designs to clarify temporal relationships between bipolar disorder, circadian disruption, and metabolic dysfunction.

## Conclusion

We found no evidence that the association between bipolar disorder and type 2 diabetes differed by chronotype, either in the overall sample or after stratification by sex. Future studies could apply objective, prospective measures of circadian function to better understand how circadian disruption interacts with bipolar disorder over time to influence cardiometabolic outcomes.

## Data Availability

We analysed third-part data for this paper that we do not have the rights to share.
Data is owned by Our Future Health (OFH), a UK-based charitable organisation (https://ourfuturehealth.org.uk/). OFH is a collaborative effort involving public sector organisations, life science companies and health charities It is funded by UK Research and Innovation (UKRI).
Data can be accessed through a Trusted Research Environment (TRE) supplied by DNAnexus as outlined on the OFH website (https://research.ourfuturehealth.org.uk/apply-to-access-the-data/) by authorised researchers named on specific projects approved by the OFH Access Board. Applications to the OFH Access Board can be made through the OFH website (https://ourfuturehealth.org.uk/about-us/how-were-governed/the-access-board/).

https://github.com/DSwiffen/OFH-ChronotypeDiabetesBipolar-August2026.git

https://ourfuturehealth.org.uk/about-us/how-were-governed/the-access-board/

## Acknowledgements

This study uses de-identified data held by Our Future Health. We acknowledge the research participants who have donated their data to the Our Future Health research programme.

This study was supported by funding from the UK Medical Research Council (MR/Z503563/1) and the Galashan Trust.

## Author contributions

DS: Conceptualisation, methodology, formal analysis, visualisation, writing (original draft), writing (review & editing); AMRL: methodology, validation, writing (review & editing); CS: methodology, writing (review & editing); HD: methodology, writing (review & editing); KF: methodology, writing (review & editing); CW: methodology, writing (review & editing), supervision; DJS: methodology, funding acquisition, writing (review & editing), supervision.

## Data statement

We analysed third-part data for this paper that we do not have the rights to share.

Data is owned by Our Future Health (OFH), a UK-based charitable organisation (https://ourfuturehealth.org.uk/). OFH is a collaborative effort involving public sector organisations, life science companies and health charities It is funded by UK Research and Innovation (UKRI).

Data can be accessed through a Trusted Research Environment (TRE) supplied by DNAnexus as outlined on the OFH website (https://research.ourfuturehealth.org.uk/apply-to-access-the-data/) by authorised researchers named on specific projects approved by the OFH Access Board. Applications to the OFH Access Board can be made through the OFH website (https://ourfuturehealth.org.uk/about-us/how-were-governed/the-access-board/).

