## supplementary_figure_1 for "Does chronotype influence the odds of diabetes comorbidity in bipolar disorder? A cross-sectional analysis of the Our Future Health cohort"

### Supplementary Figure 1: Flow chart of participants

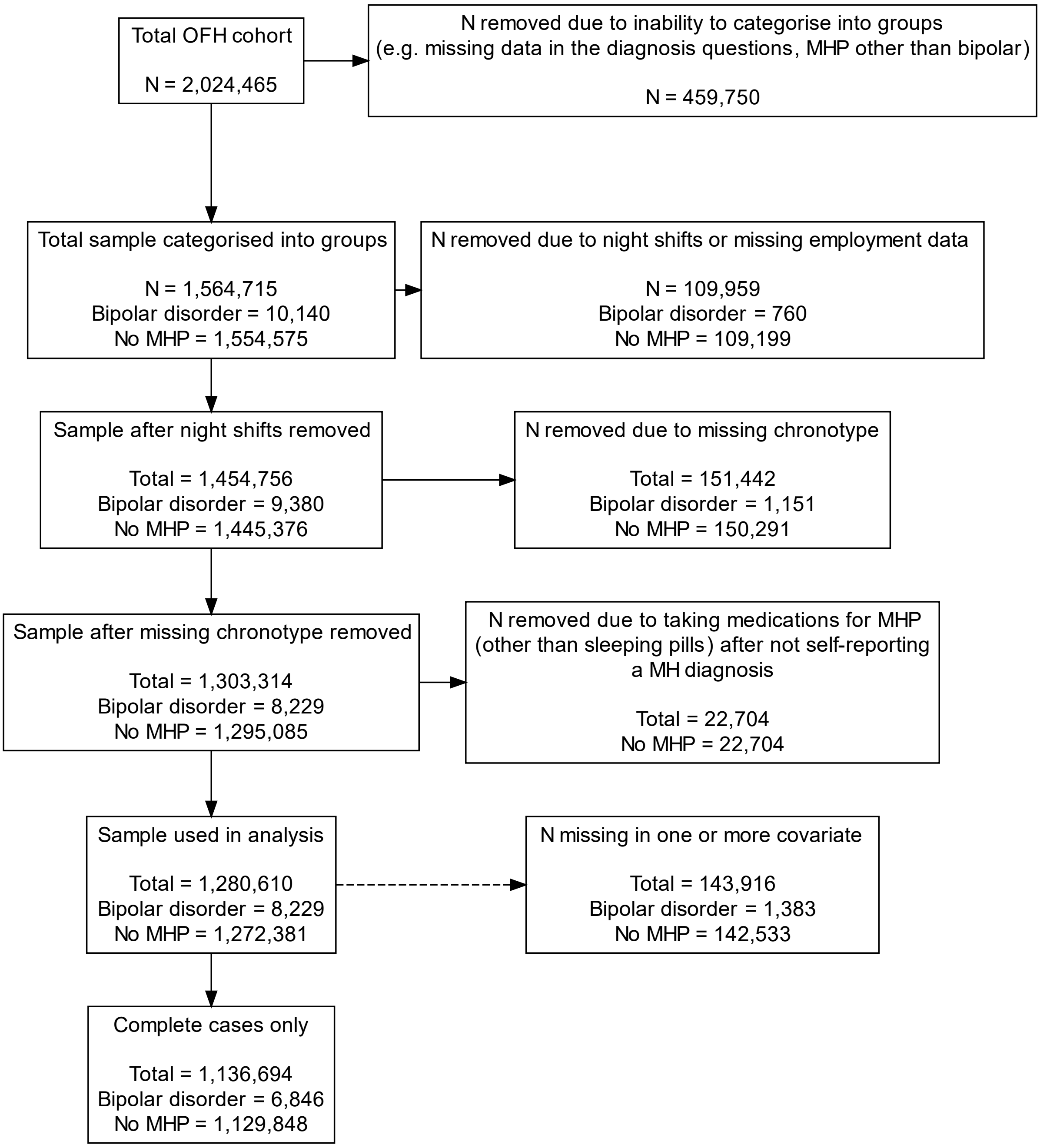


Flow chart to show the number of participants excluded from analysis and the reasons for exclusion. OFH, Our Future Health; N, sample size; MHP, mental health problem; MH, mental health
