## supplementary_methods for "Does chronotype influence the odds of diabetes comorbidity in bipolar disorder? A cross-sectional analysis of the Our Future Health cohort"

**Mental health groups**

We categorised participants into two mental health status groups: i) Bipolar disorder, and ii) No mental health problems, based on answers given to two questions in the baseline questionnaire. The first question – "*Have you ever been diagnosed with any of the following by a doctor or other health professional*?" – is offered alongside a list of body systems disorders (e.g. *Autoimmune disorder, Cancer, Neurological disorders*, etc.), and those who answered *Mental Health Problems* are then offered the second question – "*Have you ever been diagnosed with one of the following, even if you don't have it currently*?" – alongside a list of more specific psychiatric diagnoses, including bipolar disorder. Those who selected bipolar disorder (regardless of whether they selected multiple diagnoses) made up the first group.

We created the second group – No mental health problems – from participants who had given a valid response to the first question, not including those who selected *Mental Health Problems*. In this way, we removed all participants who self-reported mental health diagnoses other than bipolar. Participants who therefore answered the first question by selecting any of the other body systems disorders listed or who answered *None of the above* were included in this group. We removed those who answered *Do not know*, *Prefer not to answer* and those who provided no answer to this question from further analysis along with those who provided the answers *Do not know, Prefer not to answer,* or *None of the above* to the second question as these were not considered valid responses. We also removed a small number of participants who had not self-reported a mental health diagnosis but who had reported taking medications for their mental health (other than sleeping medication).

We also removed participants who reported any form of night shift working (i.e. those who answered *Always, Usually* or *Sometimes* to the question "*Does your work involve night shifts?"*) from further analysis.

**Outcome variable: Type 2 diabetes**

*Type 2 diabetes* is an option available in the baseline questionnaire after selecting *Endocrine disorders* in answer to the question "*Have you ever been diagnosed with any of the following by a doctor or other health professional*?". We defined the absence of type 2 diabetes as any valid response to this question other than *Type 2 diabetes*.

**Predictor variable: Chronotype**

We generated four chronotype levels – 'Definite Morning', 'More Morning than Evening', 'More Evening than Morning' and 'Definite Evening' – from the baseline question "*Do you consider yourself to be…"* with the available options: i) *Definitely a 'morning' person,* ii) *More a 'morning' than 'evening' person*, iii) *More an 'evening' than 'morning' person*, and iv) *Definitely an 'evening' person*. Those who gave one of the alternative responses – *Do not know* or *Prefer not the answer* – were removed from further analysis.

**Covariates**

We previously described how we generated age, sex, ethnicity, smoking status, current alcohol use, educational attainment and physical activity levels variables (Swiffen et al., preprint).

IMD deciles as a marker of socioeconomic status were derived from LSOA data for participants in England using the English Indices of Deprivation 2025 available from the UK Government (Ministry of Housing Communities & Local Government, 2019).

We calculated BMI based on clinic-based measurements of height and weight for those participants who had attended an OFH clinic appointment. In order to minimise missing data, for those from whom clinic-based measurements were not available, we instead used self-reported height and weight provided through the baseline questionnaire.

We created the variable for the use of sleeping medications through two baseline questions: i) "*Do you regularly take medications for any of the following reasons?*", which is offered alongside a list of body system disorders including *Mental Health Problems*, and ii) "*Do you regularly take any of the following medications…*", of which *Sleeping pills (e.g. Antihistamines, melatonin, zopiclone, barbiturates)* is an available response. We categorised all participants who selected this response into one group and into the second group we categorised all those who provided a valid answer to the first question as well as those who selected a psychotropic medication other than sleeping pills.

We created the physical multimorbidity variable using the baseline question "*Have you ever been diagnosed with any of the following by a doctor or other health professional*?". We used all valid responses to any of the physical health systems disorders (i.e. excluding *Mental Health Problems*), including *Endocrine disorders* other than type 2 diabetes. We grouped participants into three categories: 'Zero physical comorbidities', '1-2 physical comorbidities', and '≥3 physical comorbidities'.
