## supplementary_table_1 for "Does chronotype influence the odds of diabetes comorbidity in bipolar disorder? A cross-sectional analysis of the Our Future Health cohort"

| Variable | Participants with complete data (*N* = 1,136,694) | | | | Participants with incomplete data* (*N* = 143,916) | | | |
|---|---|---|---|---|---|---|---|---|
| | Bipolar disorder (*N* = 6,846) | | No mental health problems (*N* = 1,129,848) | | Bipolar disorder (*N* = 1,383) | | No mental health problems (*N* = 142,533) | |
| **Female sex,** N (%) | 4,482 | (65.5) | 618,779 | (54.8) | 960 | (69.4) | 83,865 | (58.8) |
| **Age**, median years [IQR] | 50 | [37, 61] | 57 | [43, 67] | 45 | [34, 58] | 57 | [41, 67] |
| **Ethnicity** | | | | | | | | |
| Black, N (%) | 62 | (0.9) | 12,985 | (1.1) | 14 | (1.0) | 2,064 | (1.4) |
| Mixed or multiple heritage ethnic background, N (%) | 132 | (1.9) | 12,786 | (1.1) | 16 | (1.2) | 1,544 | (1.1) |
| Other, N (%) | 198 | (2.9) | 30,294 | (2.7) | 41 | (3.0) | 4,031 | (2.8) |
| South Asian, N (%) | 140 | (2.0) | 43,076 | (3.8) | 34 | (2.5) | 5,462 | (3.8) |
| White, N (%) | 6,314 | (92.2) | 1,030,707 | (91.3) | 1,267 | (91.6) | 127,615 | (89.5) |
| **Type 2 diabetes, N (%)** | 452 | (6.6) | 25,531 | (2.3) | 76 | (5.5) | 3,112 | (2.2) |
| **Chronotype** | | | | | | | | |
| Definite morning, N (%) | 1,230 | (18.0) | 297,776 | (26.4) | 226 | (16.3) | 37,945 | (26.6) |
| More morning than evening, N (%) | 1,736 | (25.4) | 415,930 | (36.8) | 298 | (21.5) | 49,294 | (34.6) |
| More evening than morning, N (%) | 2,177 | (31.8) | 305,592 | (27.0) | 469 | (33.9) | 39,746 | (27.9) |
| Definite evening, N (%) | 1,703 | (24.9) | 110,550 | (9.8) | 390 | (28.2) | 15,548 | (10.9) |
| **Smoking:** History of regular cigarette, cigar or tobacco pipe smoking, N (%) | 2,220 | (32.4) | 314,229 | (27.8) | 414 | (29.9) | 32,102 | (22.5) |
| **Alcohol** | | | | | | | | |
| Drinks alcohol more frequently than once or twice per week, N (%) | 1,427 | (20.8) | 349,264 | (30.9) | 199 | (14.4) | 35,676 | (25.0) |
| Drinks alcohol once or twice a week or less frequently, N (%) | 3,950 | (57.7) | 673,539 | (59.6) | 810 | (58.6) | 88,410 | (62.0) |
| Never drinks alcohol, N (%) | 1,469 | (21.5) | 107,045 | (9.5) | 339 | (24.5) | 16,580 | (11.6) |
| **Physical activity levels** | | | | | | | | |
| High level physical activity, N (%) | 18 | (0.3) | 36.5 | (0.3) | NR | | 505 | (0.4) |
| Low to medium level physical activity, N (%) | 5,709 | (83.4) | 1,073,063 | (95.0) | 1047 | (75.7) | 129,603 | (90.9) |
| No physical activity, N (%) | 1,119 | (16.3) | 53,180 | (4.7) | NR | | 10,225 | (7.2) |

| | | | | | | | | |
|---|---|---|---|---|---|---|---|---|
| **Body mass index**, median kg/m$^2$ [IQR] | 28 | [25, 33] | 26 | [23, 29] | 28 | [24, 33] | 26 | [24, 30] |
| **College or University degree**, N (%) | 3,518 | (51.4) | 608,388 | (53.8) | 624 | (45.1) | 67,670 | (47.5) |
| **Multiple Deprivation Index**, median decile [IQR] | | | | | | | | |
| IMD quintile 1 (most deprived), N (%) | 1,520 | (22.2) | 104,575 | (9.3) | 223 | (16.8) | 10,406 | (7.3) |
| IMD quintile 2, N (%) | 1,469 | (21.5) | 169,093 | (15.0) | 175 | (12.7) | 11,937 | (8.4) |
| IMD quintile 3, N (%) | 1,355 | (19.8) | 223,677 | (19.8) | 110 | (8.0) | 13,004 | (9.1) |
| IMD quintile 4, N (%) | 1,280 | (18.7) | 282,734 | (25.0) | 114 | (8.2) | 14,225 | (10.0) |
| IMD quintile 5 (least deprived), N (%) | 1,222 | (17.8) | 349,769 | (31.0) | 98 | (7.1) | 15,521 | (10.9) |
| **Taking sleeping medications**, N (%) | 985 | (14.4) | 706 | (0.1) | 257 | (18.6) | 100 | (0.1) |
| **Physical comorbidities** | | | | | | | | |
| 0, N (%) | 1,485 | (21.7) | 402,864 | (35.7) | 283 | (20.5) | 51,867 | (36.4) |
| 1-2, N (%) | 2,864 | (41.8) | 562,976 | (49.8) | 566 | (40.9) | 69,571 | (48.8) |
| ≥3, N (%) | 2,497 | (36.5) | 164,008 | (14.5) | 505 | (36.5) | 19,921 | (14.0) |

**Baseline characteristics of complete versus incomplete cases separated by diagnostic group**

Sociodemographic, lifestyle, socioeconomic and health-related factors are shown for both the complete and incomplete cases across both groups of participants with self-reported bipolar disorder and participants with no self-reported mental health problems. IQR, interquartile range; LSOA, Lower Layer Super Output Areas; NR, not reported due to <10 observations in one cell per covariate. As totals are given, cells with the second smallest number of observations per covariate were also redacted to prevent back calculation and risk of disclosure.

* For each covariate under "Participants with incomplete data", the percentage reported is based on participants who did not have missing data for that covariate.
